# Rates of growth during and after nutritional rehabilitation and liver fat in adult survivors of severe acute malnutrition

**DOI:** 10.1101/2022.06.08.22276056

**Authors:** Debbie Thompson, Kimberley McKenzie, Asha Badaloo, Carolyn Taylor-Bryan, Ingrid Tennant, Deanne Soares, Terrence Forrester, Michael Boyne

**Affiliations:** Caribbean Institute for Health Research, The University of the West Indies, Kingston, Jamaica; Department of Surgery, Radiology, Anesthesia and Intensive Care, The University of the West Indies, Kingston, Jamaica; UWI Solutions for Developing Countries, The University of the West Indies, Kingston, Jamaica; Department of Medicine, The University of the West Indies, Kingston, Jamaica

**Keywords:** Non-alcoholic fatty liver disease, rehabilitation growth, marasmus, kwashiorkor

## Abstract

**Background:** Nutritional rehabilitation during severe malnutrition (SM) aims to quickly restore a healthy body weight, but rapid weight gain has been associated with later cardiovascular risk. We hypothesized that faster weight gain during SM rehabilitation and post-hospitalization is associated with liver fat in adult survivors.

**Method:** Adult survivors of childhood SM underwent abdominal CT scan to estimate liver fat as mean liver attenuation (MLA) and liver spleen ratio (L/S). Birth weight (BW) and anthropometry measured during, and post-hospitalization were abstracted from admission records.

**Results:** We studied 42 marasmus survivors (Ms) and 40 kwashiorkor survivors (Ks). Ms had a lower mean BW (SD) 2.5 (0.8) vs 3.0 (0.7) kg; *p*=0.01) and were more wasted (*p*<0.001) and stunted (*p*=0.03) than Ks on admission to hospital. Ms and Ks had similar rates of rehabilitation weight gain, which was inversely associated with MLA among all survivors of SM (r=-0.246, *p*=0.029), but only in Ms when assessed by diagnosis (r= -0.449, *p*=0.004). The association between rehabilitation weight gain and adult liver fat in Ms was not altered by BW, admission wasting or stunting. In Ks, post-hospitalization height gain was inversely associated with MLA (difference = -0.64, 95%CI: -0.64 to -0.13; *p*=0.006).

**Conclusions:** Faster rehabilitation weight gain is associated with liver fat in adult survivors of childhood severe malnutrition. The finding that birth weight did not influence these outcomes may reflect the timing of the nutritional insult in utero. Target weight gain during nutritional rehabilitation may need to be lowered to optimize long-term outcomes in these children.

## Introduction

Early life exposure to severe malnutrition has been reported to be associated with later non-communicable diseases [1, 2] possibly including non-alcoholic fatty liver disease (NAFLD). Indeed, famine exposure in early life has been shown to have a significant association with the risk of NAFLD in adult women [3]. Factors that might influence liver fat accumulation in persons fully recovered from severe malnutrition are birth weight, the degree of wasting and/or stunting on admission and the rate of weight gain during nutritional rehabilitation [4, 5]. Additionally, the propensity to accumulate fat in the liver might differ between the oedematous (kwashiorkor) and non-oedematous (marasmus) forms of malnutrition as we previously demonstrated that adult survivors of marasmus (Ms) have more liver fat than adult survivors of kwashiorkor (Ks) [6]. However, the reason for this finding is not clear.

Several studies report that the rate and total weight gain during nutritional rehabilitation are greater influences on the development of fatty liver than the initial nutritional insult. Faienza et al compared 23 children who were small for gestational age (SGA) and experienced rapid weight gain within the first 6-12 months to 24 children whose birth weights were appropriate for gestational age (AGA) [7]. NAFLD as measured by abdominal ultrasonography, was observed in 35% of SGA children and was cited as an emerging problem in SGA pre-pubertal children who experienced a rapid weight gain in postnatal life [7]. Additionally, data from a South-west England population-based birth cohort reported that greater gains in weight-for-age between years 1 and 10 years are associated with liver fat in adolescence (by ultrasonography) in children who were not previously malnourished [8].

Established inpatient management of SAM begins with an acute “stabilisation” phase in which a diet of 80-100kcal/kg/day is fed to provide energy for weight maintenance, while infections and fluid imbalance are treated [9]. Once there is clinical improvement, loss of all oedema and improved appetite and affect, the “rehabilitation” phase begins during which children are fed increasing amounts of energy and protein enriched feeds to allow for attainment of a healthy body weight. The WHO recommends that optimal weight gain during this period is >10 g/kg/day [10]. The final phase of treatment, referred to as “recovery”, involves weaning the child to an age-appropriate diet before discharge from hospital.

Jamaican children with marasmus are reported to weigh less at birth [11], and are more wasted and stunted on admission to hospital [12] than children with oedematous malnutrition. We hypothesize that greater weight gains during rehabilitation and post-hospitalization are associated with greater liver fat in adult survivors of SAM. We also hypothesized that, compared to children with oedematous malnutrition, children with marasmus would have more liver fat as adults due to their lower birth weight. This study therefore investigated the relationships between gains in weight and height (during and immediately after SAM rehabilitation) and adult liver fat.

## Methods

### Study Design/ Subjects

This was a cross-sectional study of 88 adults who were admitted during early childhood to the Tropical Metabolism Research Unit (TMRU) Ward with a diagnosis of severe malnutrition. A retrospective cohort was assembled by reviewing ward records of patients admitted to the TMRU of the University of the West Indies between 1963 and 1993. Using the Wellcome criteria, patients were classified as having marasmus (weight-for-age < 60%) or kwashiorkor (60-80% weight-forage, plus the presence of oedema) in comparison to the National Centre for Health Statistics (NCHS) standard growth curves [13]. Of 1,336 patients admitted during the period, 729 individuals were traced, 316 had basic anthropometric measurements [11] and 92 underwent abdominal CT scans (**Figure 1**).

**Figure 1:**
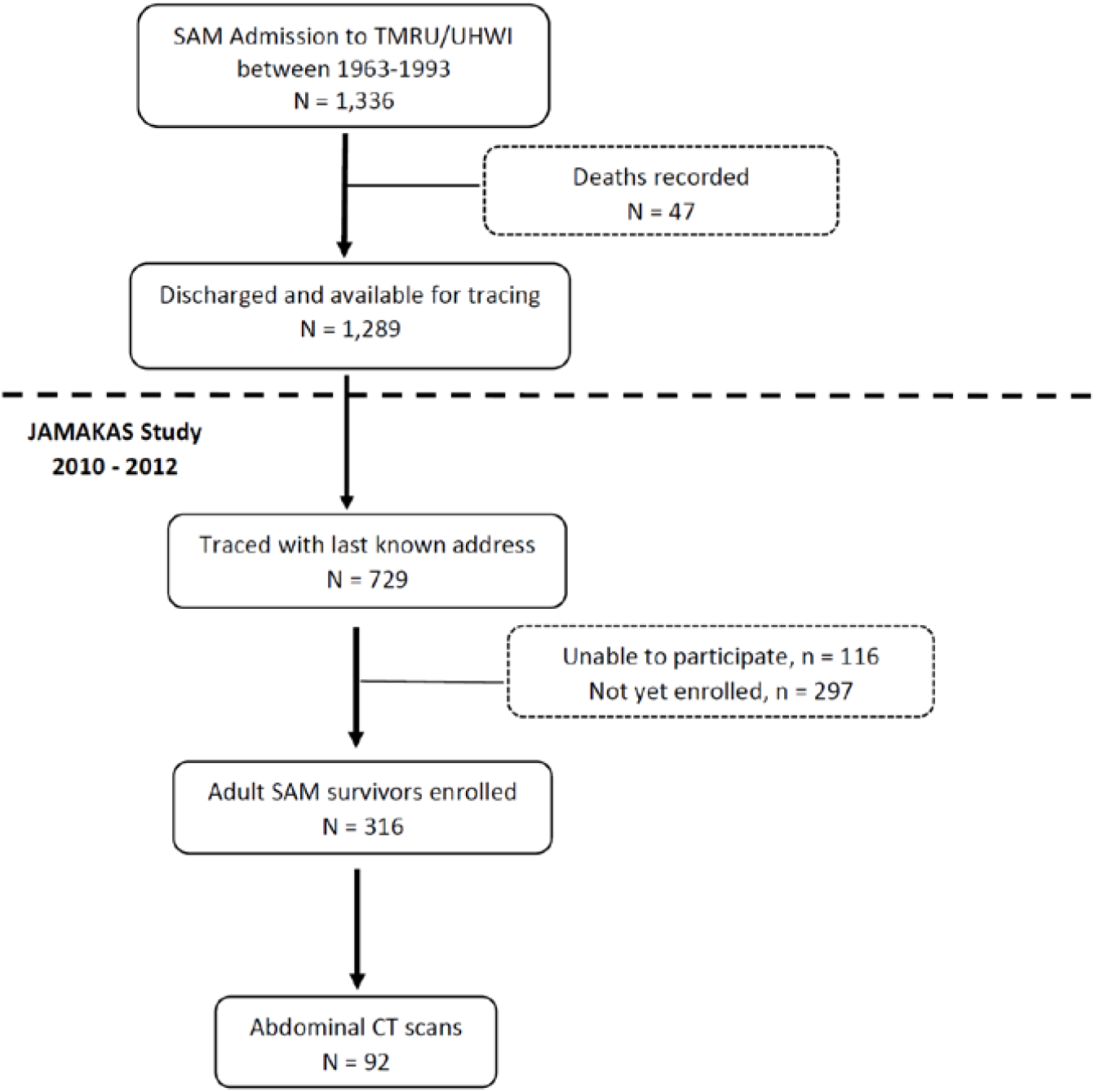
Flow chart detailing recruitment of adult survivors of SAM (*n* = 92). “Unable to participate” includes adult survivors of SAM who were unavailable because of migration (*n* = 53), illness (*n* = 19), refusal (*n* = 14), or pregnancy (*n* = 30). TMRU, Tropical Metabolism Research Unit; UHWI, University Hospital of the West Indies; JAMAKAS, Jamaica Marasmus and Kwashiorkor Adult Survivors.

Persons with sickle cell haemoglobinopathy, a history of liver disease or taking medications that cause hepatic abnormalities were excluded from the study and participants with a self-reported alcohol intake of more than 14 drinks per week (males) and more than 7 drinks per week (females) [14] were excluded from the analyses.

The Mona Campus Research Ethics Committee of the University of the West Indies approved the study protocol (ECP17, 14/15), and each participant gave written informed consent. Written consent was also obtained at the time of admission to hospital (as children) for medical records to be used for research.

### Paediatric Measurements

Recalled birth weights and height and weight data collected at and during hospitalization were abstracted from admission records. As children were followed up for a minimum of 2 years upon discharge from hospital, height and weight collected over this period were also available for abstraction. The inpatient measurements were taken by research nurses trained in the measurement of length/height and weight. Length was measured weekly using an infantometer and weight daily using a digital scale. Outpatient measurements were taken either at the TMRU clinic or during a home visit. In the latter case a portable stadiometer (Seca 213) and scale were used.

Rates of weight gain were calculated over 2 time periods as follows:

1. During the rehabilitation phase i.e., the weight difference between weights recorded from the beginning of rehabilitation feeds (when all oedema had resolved) to the end of rehabilitation phase).
2. In the post-hospitalization period (discharge weight to final follow-up weight).

Rates of height gain were calculated over 2 time periods as follows:

1. During rehabilitation phase i.e., height recorded from the beginning of rehabilitation feeds (when all oedema had resolved) to the end of rehabilitation phase.
2. In the post-hospitalization period (discharge height to final follow up height). Where there was no height recorded on discharge, the last height recorded before discharge from hospital was used.

### Adult Measurements

After a 10-hour overnight fast, participants reported to the TMRU where they completed a staff-administered questionnaire. Body weight was measured to the nearest 0.1 kg and height and waist circumference to the nearest 0.1cm using a standardized protocol [15].

### Liver Fat

Abdominal CT scans (Phillips Brilliance 64-slice scanner) were conducted to measure liver fat. A 5mm CT scan was taken at the T12/L1 intervertebral disc space to include both the liver and the spleen. Using eFilm Workstation 3.1 (Merge Healthcare, Chicago, IL, USA), three regions of interest were placed in the liver and one in the spleen and attenuation readings taken. A mean liver attenuation (MLA) of < 40HU and liver spleen: ratio (L/S) <1 denoted hepatic steatosis > 30%.

### Statistical Analysis

SPSS 19.0 for Windows was used for the statistical analyses. Characteristics of the survivors of marasmus and kwashiorkor were compared using the independent T test. Multivariate regression analyses were conducted with models adjusting for sex, and adult age, BMI / fat mass, as all are documented to be associated with fatty liver. Two-sided p-values were reported and a *p*-value ≤ 0.05 considered statistically significant.

## Results

Hospital records were available for 82 of the 92 participants who underwent abdominal CT scan **(Figure 1)**.

The mean (SD) age of the adult survivors was 28.8 (8.5) years, BMI was 23.8 (5.6) kg/m^2^, MLA was 64.0 (4.7) HU and liver: spleen ratio (L/S) was 1.3 (0.19). Of the 82 SAM survivors, 42 were diagnosed with marasmus and 40 with kwashiorkor as children. The participants’ mean (SD) birth weight was 2.7 (0.8) kg, and on admission: age, 11.7 (4.6) months, weight, 5.6 (1.4) kg, height, 65.3 (6.4) cm, WHO weight-for-age z-scores, -4.05 (1.41) **(Table 1)**. The mean (SD) rate of weight gain during rehabilitation was 9.3 (3.6) g/kg/day and the mean (SD) height gain during the same period was 1.3 (1.5) cm (**Table 1**).

**Table 1:** Summary of birth weight, admission anthropometry and gains in weight and height during and after hospital admission for SAM.

|  | All Participants<br>(N=82) | Marasmus<br>(N=42) | Kwashiorkor<br>(N=40) | P-value<br>(Ms vs. Ks) |
| --- | --- | --- | --- | --- |
| Birth Weight (kg) | 2.7 ± 0.8 | 2.5 ± 0.8 | 3.0 ± 0.7 | 0.01 |
| **Admission** |  |  |  |  |
| Age (days) | 356 ± 139 | 364.2 ± 150.0 | 347.8 ± 128.7 | 0.60 |
| Sex (% male) | 52% | 55% | 50% |  |
| Weight (kg) | 5.6 ± 1.4 | 5.1 ± 1.2 | 6.3 ± 1.3 | <0.001 |
| Height/ Length (cm) | 65.3 ± 6.4 | 64.6 ± 6.8 | 66.1 ± 5.9 | 0.32 |
| Weight-for-age z-score (WAZ) | -4.1 ± 1.4 | -4.9 ± 1.2 | -3.2 ± 1.1 | <0.001 |
| Length/height-for-age z-score (HAZ) | -3.6 ± 1.7 | -4.0 ± 1.9 | -3.2 ± 1.3 | 0.031 |
| Weight-for-length z-score (WHZ) | -2.8 ± 1.4 | -3.8 ± 0.9 | -1.8 ± 1.2 | <0.001 |
| **Rehabilitation Phase (minimum weight to maximum weight)** |  |  |  |  |
| Absolute weight gain (kg) | 1.9 ± 0.9 | 2.0 ± 0.8 | 1.7 ± 0.9 | 0.13 |
| Weight gain (g/d) | 57.7 ± 21.0 | 53.3 ± 17.6 | 62.3 ± 23.3 | 0.053 |
| Weight gain (g/kg/d) | 9.3 ± 3.6 | 9.2 ± 4.0 | 9.3 ± 3.2 | 0.92 |
| Change in weight-for age z-score (ΔWAZ) | 2.1 ± 0.9 | 2.2 ± 0.9 | 1.9 ± 0.9 | 0.13 |
| Absolute height gain (cm) | 1.3 ± 1.5 | 1.6 ± 1.9 | 1.0 ± 1.2 | 0.45 |
| Height gain (cm/y) | 13.0 ± 12.2 | 12.2 ± 14.0 | 13.5 ± 11.5 | 0.83 |
| Change in height-for age z-score (ΔHAZ) |  |  |  |  |
| **Post-hospitalization Phase (from discharge up to 2 years follow up)** |  |  |  |  |
| Weight gain (g/d) | 6.1 (3.2, 8.7) | 3.9 (1.0, 7.3) | 6.8 (5.2, 9.0) | 0.18 |
| Weight gain (g/kg/month) | 17.0 ± 26.9 | 14.2 ± 18.2 | 19.9 ± 33.7 | 0.39 |
| Change in WAZ/month (ΔWAZ) | -0.02 ± 2.2 | -0.004 ± 0.14 | 0.005 ± 0.27 | 0.43 |
| Height gain from last height during admission (cm/y) | 13.5 ± 5.9 | 11.7 ± 3.5 | 15.4 ± 7.4 | 0.08 |
| Height gain from 1<sup>st</sup> discharge height (cm/y) | 14.0 ± 6.8 | 13.0 ± 5.5 | 14.9 ± 7.7 | 0.37 |
| Change in HAZ/month (ΔHAZ) | 0.1 ± 0.16 | 0.11 ± 0.18 | 0.09 ± 0.12 | 0.63 |
| **Adult Anthropometry, Body Composition and Liver Fat** |  |  |  |  |
| Age (years) | 28.8 ± 8.5 | 26.7 ± 7.6 | 31.1 ± 8.9 | 0.02 |
| BMI (kg/m<sup>2</sup>) | 23.8 ± 5.6 | 22.4 ± 5.5 | 25.3 ± 5.4 | 0.02 |
| Lean Mass (kg) | 46.9 ± 9.4 | 45.7 ± 9.0 | 48.3 ± 9.7 | 0.21 |
| Fat Mass (kg) | 15.9 ± 13.7 | 13.3 ± 13.1 | 18.5 ± 13.9 | 0.09 |
| MLA (HU) | 64.0 ± 4.7 | 63.5 ± 4.4 | 64.6 ± 5.0 | 0.29 |
| L/S | 1.3 ± 0.19 | 1.2 ± 0.2 | 1.3 ± 0.2 | 0.65 |

Ms had a lower birth weight, and, on admission to hospital, they weighed less and were lighter using weight-for-age z-scores (WAZ), more stunted using height-for-age z-scores (HAZ) and more wasted using weight-for-length z-scores (WLZ) than Ks (**Table 1**). Rates of rehabilitation weight gain were similar in Ms and Ks using g/d (*p* = 0.053), g/kg/day (*p* = 0.69) and change in WAZ (ΔWAZ) (*p* = 0.13) (**Table 1**). As adults, Ks were older than Ms and had a higher BMI (*p* = 0.02) (**Table 1**).

Among these adult survivors of severe malnutrition, the rate of weight gain during rehabilitation (in g/d) was inversely associated with MLA adjusting for age and sex (r = -0.247, *p* = 0.027), and after additionally adjusting for adult BMI (r = -0.246, *p* = 0.028) (**Figure 2**) or fat mass (r = - 0.247, *p* = 0.028). Rehabilitation weight gain expressed as g/kg/day was inversely associated with MLA adjusting for age and sex (r = -0.224; *p* = 0.045) and after additionally adjusting for fat mass (*p* = 0.05) but not after adjusting for BMI (*p* = 0.053). This effect of rehabilitation weight gain (in g/kg/day) on MLA did not depend on the value of adult BMI (*p* = 0.71) However, rehabilitation weight gain expressed as ΔWAZ was not related to MLA (*p* = 0.28). Height gain during rehabilitation was not related to MLA or L/S (*p* > 0.6). Post-hospitalization weight and height gain were not associated with MLA or L/S (*p* ≥ 0.052) (data not shown).

**Figure 2:**
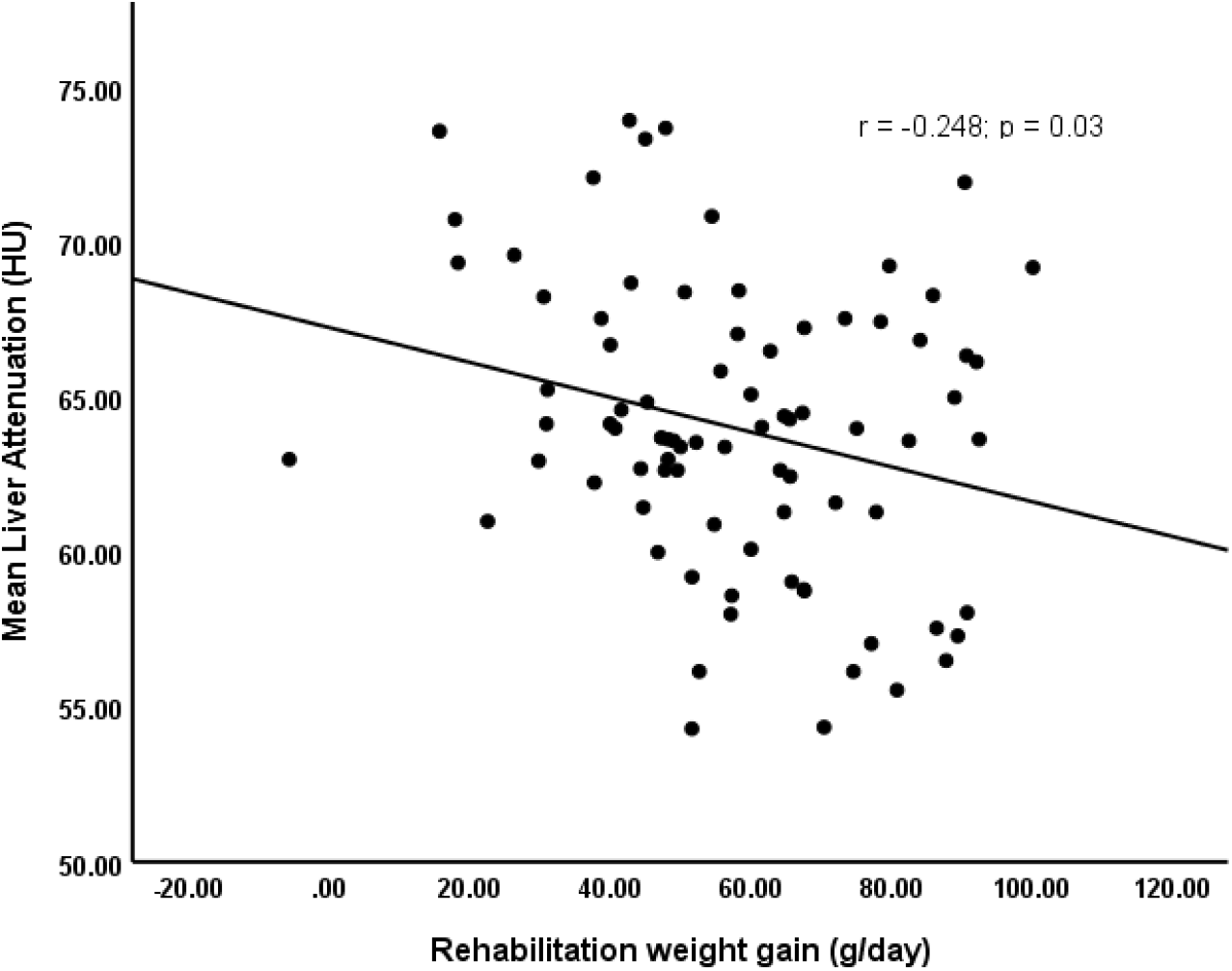
Correlation between mean liver attenuation and rate of rehabilitation weight gain (age, sex and BMI-adjusted) in adult survivors of SAM.

Effect of diagnosis: When the participants were grouped by diagnosis (i.e., Ms and Ks), and regression analyses were adjusted for age, sex and BMI, faster rehabilitation weight gain (g/d) had a negative association with both MLA (difference = -0.11 ± 0.04, *p* = 0.003) (**Table 2**) and L/S (difference =-0.004 ± 0.002, *p* = 0.024) in Ms. However, among these marasmus survivors, MLA was not associated with rehabilitation weight gain expressed in g/kg/day (*p* = 0.15) or change in WAZ (*p* = 0.25); the same is true for LS (*p* > 0.3). In Ks, rehabilitation weight gain was not associated with MLA in a regression model adjusting for age, sex and adult BMI. However, height gain post-discharge (cm/year) was negatively associated with MLA in Ks (difference = -0.384 + 0.12, *p* = 0.006) (**Table 2**).

**Table 2:**
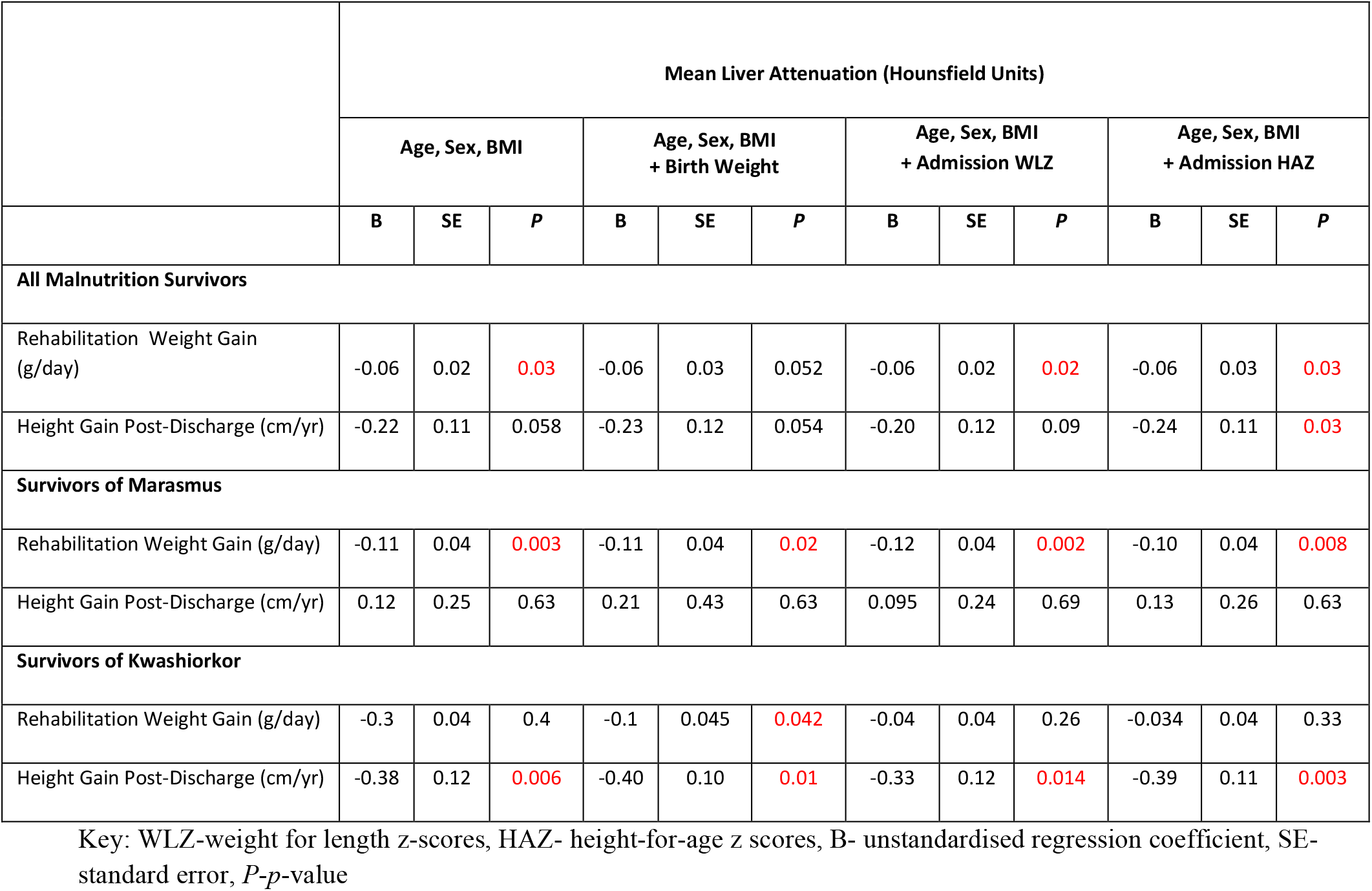
Multivariate age-and-sex-adjusted regression analyses of selected growth variables against mean liver attenuation by diagnosis of SAM (models shown)

|  | Mean Liver Attenuation (Hounsfield Units) |  |  |  |  |  |  |  |  |  |  |  |
| --- | --- | --- | --- | --- | --- | --- | --- | --- | --- | --- | --- | --- |
|  | Age, Sex, BMI |  |  | Age, Sex, BMI + Birth Weight |  |  | Age, Sex, BMI + Admission WLZ |  |  | Age, Sex, BMI + Admission HAZ |  |  |
|  | B | SE | P | B | SE | P | B | SE | P | B | SE | P |
| **All Malnutrition Survivors** |  |  |  |  |  |  |  |  |  |  |  |  |
| Rehabilitation Weight Gain (g/day) | -0.06 | 0.02 | 0.03 | -0.06 | 0.03 | 0.052 | -0.06 | 0.02 | 0.02 | -0.06 | 0.03 | 0.03 |
| Height Gain Post-Discharge (cm/yr) | -0.22 | 0.11 | 0.058 | -0.23 | 0.12 | 0.054 | -0.20 | 0.12 | 0.09 | -0.24 | 0.11 | 0.03 |
| **Survivors of Marasmus** |  |  |  |  |  |  |  |  |  |  |  |  |
| Rehabilitation Weight Gain (g/day) | -0.11 | 0.04 | 0.003 | -0.11 | 0.04 | 0.02 | -0.12 | 0.04 | 0.002 | -0.10 | 0.04 | 0.008 |
| Height Gain Post-Discharge (cm/yr) | 0.12 | 0.25 | 0.63 | 0.21 | 0.43 | 0.63 | 0.095 | 0.24 | 0.69 | 0.13 | 0.26 | 0.63 |
| **Survivors of Kwashiorkor** |  |  |  |  |  |  |  |  |  |  |  |  |
| Rehabilitation Weight Gain (g/day) | -0.3 | 0.04 | 0.4 | -0.1 | 0.045 | 0.042 | -0.04 | 0.04 | 0.26 | -0.034 | 0.04 | 0.33 |
| Height Gain Post-Discharge (cm/yr) | -0.38 | 0.12 | 0.006 | -0.40 | 0.10 | 0.01 | -0.33 | 0.12 | 0.014 | -0.39 | 0.11 | 0.003 |

Key: WLZ-weight for length z-scores, HAZ- height-for-age z scores, B- unstandardised regression coefficient, SE- standard error, P-p-value

Effect of birth weight: In these participants, birth weight was not associated with adult liver fat (*p* = 0.10) adjusting for age, sex and BMI. After additionally adjusting the model for birth weight, the relationship between rehabilitation weight gain and MLA observed in Ms (*p* = 0.021) (**Table 2**) remained significant, but the inverse association between rehabilitation weight gain (g/day) and MLA seen in all the participants was no longer significant (*p* = 0.052). The interaction term (birth weight and rehabilitation weight gain) was not significant (*p* = 0.39). In Ks, although adjusting for birth weight did not affect the associations between post-hospitalization height gain and MLA, it revealed an association between rehabilitation weight gain and MLA **(Table 2)**.

Effect of admission HAZ: Adjusting for age, sex, BMI and admission HAZ, there was an inverse association between rate of rehabilitation weight gain and mean liver attenuation (*p* = 0.03) among all the participants, and this relationship was also significant in Ms (*p* = 0.008). In Ks, the inverse correlation between rate of height gain and mean liver attenuation remained significant when the age, sex and BMI-adjusted regression was further adjusted for HAZ (*p* = 0.003) (**Table 2**).

Effect of admission WLZ: In general, WLZ on admission had no relationship with either outcome measure of liver fat adjusting for age, sex and BMI (*p-*values > 0.7). In Ms, adding admission WLZ to the age, sex and BMI-adjusted regression model did not affect the inverse association between rate of rehabilitation weight gain and MLA (*p* = 0.002). Similarly, adjusting for admission WLZ on admission in Ks did not alter the inverse relationship between height gain and MLA (*p* = 0.014) (**Table 2**).

Effect of admission WAZ: Admission WAZ was assciated with rehabilitation weight gain (g/day) even after adjusting for age and sex (r = 0.25; *p* = 0.025). Despite this, the inverse association between rehabilitation weight gain (g/day) and MLA seen in all the malnutrition survivors remained significant (*p* = 0.03) after adjusting for admission WAZ (data not shown).

## Discussion

This study investigated the associations between weight and height gain during and post-hospitalization for SAM and later liver fat in an Afro-Caribbean population. Our findings supported our hypothesis that, among survivors of malnutrition, there is a direct association between childhood gains in weight during rehabilitation (in g/day) and adult liver fat. This association between rehabilitation weight gain and adult liver fat accumulation was also specifically seen in marasmus survivors. We also demonstrated a direct relationship between post-hospitalization height gain (cm/year) and adult liver fat in kwashiorkor survivors only.

It has been reported that, during nutritional rehabilitation, children in the lowest WHZ tertile tend to have greater absolute weight gain, faster weight gain (g/kg/day) and higher increase in WHZ score at 1 month when compared to the children in the highest tertile of WHZ [16]. However, although admission WAZ was associated with faster rehabilitation weight gain in g/day in our participants, WLZ on admission was not. Despite this, the positive association between rehabilitation weight gain and adult liver fat was not influenced by neither admission weight, admission weight-for-age z-score or minimum weight in our participants. The WHO recommends a vigorous approach to feeding during the rehabilitation phase, aimed at achieving weight gain of >10 g /kg/d [10]. Our participants, with a slightly lower mean rehabilitation weight gain of 9.3 g/kg/day, already appear to be at greater risk of fatty liver. As this association between rehabilitation weight gain (in g/kg/d) and adult liver fat is influenced by adult BMI, it will be important to optimize adult body size in these SAM survivors to reduce the risk of liver fat accumulation.

We demonstrated distinct associations with adult liver fat by diagnosis (i.e., weight in Ms and height in Ks). Despite kwashiorkor and marasmus survivors having similar rates of weight gain during the rehabilitation phase of SAM treatment, rehabilitation weight gain was only associated with liver fat in marasmus survivors. Given their lower birth weight, and in keeping with the thrifty phenotype hypothesis, marasmus survivors may be less well adapted to a calorie-surfeit environment than kwashiorkor survivors and might be predisposed to cardiometabolic risk later in life. Thus Ms, who were smaller on admission to hospital than Ks, might be better adapted to a lower rate of weight gain. We further posit that after a nutritional insult in utero, Ms could be predisposed to one pattern of adipogenesis (the process by which mature adipocytes develop), while Ks, with an initial normal growth trajectory in utero, are predisposed to another pattern. Following an adverse *in utero* development, fetuses who experienced intrauterine growth retardation display increased lipogenic (fatty acid synthesis and subsequent triglyceride synthesis in both liver and adipose tissue) and adipogenic (differentiation of pre-adipocytes into mature fat cells) capacity in adipocytes [17]. This developmental process of adipogenesis has been shown to occur between 14-23 weeks of gestation, after which the total number of fat lobules remains approximately constant [18]. Using mice, Bieswal et al demonstrated that the size distribution of adipocytes and adipose tissue growth were both sensitive to the type of nutrient restriction as well as the time at which early malnutrition occurred [19]. Furthermore, small Indian babies had small abdominal organs and reduced lean mass, but maintained adiposity during their intrauterine development, and the authors posit that this body composition may persist postnatally [20]. If that is so, the possibility exists that Ms, who were smaller at birth than Ks, might be predisposed to laying down fat as visceral adipose tissue. However, given the age of our participants, it is difficult to say whether this is purely an effect of the timing of the nutritional insult or the effect of accumulated insults.

Unexpectedly, the association between rehabilitation weight gain and adult liver fat was not influenced by birth weight. While adjusting for birth weight did not affect the effect of rehabilitation weight gain on adult liver fat, we acknowledge including birth weight in models comparing Ms and Ks could represent over-adjusting as Ms have lower BW than Ks. However, low birth weight, as a proxy for poor intrauterine nutrition, has long been associated with adverse long-term outcomes related to NCDs. Indeed, in a systematic review of 57 studies describing the relationships between low birth weight, rehabilitation growth and features of the metabolic syndrome [21] it was concluded that both low birth weight and rehabilitation-growth were associated with aspects of subsequent metabolic syndrome [21]. While these findings were not specific to liver fat, fatty liver disease is widely regarded by many to be associated with many of the features of metabolic syndrome [22, 23]. Since birth weight is not contributory in this cohort of SAM survivors, we posit that low birth weight in this group of participants may not reflect intrauterine undernutrition occurring during those critical windows of developmental plasticity (typically 2½ - 9 weeks) that could lead to abnormal organ development and later NCD risk.

In kwashiorkor survivors, post-hospitalization height gain was associated with adult liver fat and this association was not influenced by adult height. However, gains in height are typically associated with favorable cardiovascular outcomes in adults [24]. In support, greater adult height was associated with lower risk of NAFLD among males and females in China in a prospective cohort study of more than 35,000 participants aged 25 years or over with height measured at baseline [25]. However, there is recent evidence that suggests that gains in height can also be associated with adverse outcomes, as greater height at birth, or height gain at 0-3 months of age and 6-8 years of age all were associated with higher adult cardiovascular risk traits in 2,218 adults from a birth cohort in India [26]. While these associations were attenuated when the analyses were adjusted for BMI and adult height [26], the same was not true for our participants in whom neither height at admission nor adult height influenced the association between post-hospitalization height gain and liver fat. While there are likely other intervening factors that influence the association, it is possible that, in children who were normal weight at birth, height gain occurring during certain critical windows could represent a similar risk for cardio-metabolic disease as increased adiposity.

### Strengths and Limitations

The study utilized a well-characterized cohort and the use of CT scans allowed for objective, reproducible, quantitative data. This was supported by the availability of daily weight and weekly length/height measurements throughout hospitalization. While the lack of data on current diet was a limitation, we posit that most of these SAM survivors remained in a non-obesogenic environment as they remained lean on average. We acknowledge that, along with diet, other intervening factors might influence liver fat in these adult SAM survivors. Additionally, computed tomography is known to be a less sensitive measure of liver fat than magnetic resonance spectroscopy. Finally, the study was also limited by the use of a relatively young, lean study population in whom the prevalence of fatty liver is likely to be low.

## Conclusion

Faster rehabilitation weight gain is associated with more liver fat in Afro-Caribbean adult survivors of severe malnutrition. Rehabilitation weight and post-hospitalization height gain are positively associated with liver fat in marasmus and kwashiorkor survivors respectively. The influence of birth weight was not established in these participants and may reflect the timing of the nutritional insult in utero. Efforts at rehabilitation must therefore be prudent and guided by the evidence, specifically the recommendations of the WHO [10]. Nevertheless, our data underscores the need to re-examine malnutrition treatment guidelines and optimize rehabilitation weight gain targets in children with severe malnutrition to prevent cardiometabolic risk and fatty liver disease in later life.

## Data Availability

All data produced in the present study are available upon reasonable request to the authors

## Acknowledgements

We gratefully acknowledge the men and women who took part in the study. We also recognize Professor Clive Osmond for assisting with the revision of the manuscript.

## Author Contributions

DT, AB, MB, CT-B, IT, and TF designed the research, IT, DS, DT and KM participated in data collection and coordinating the clinical samples. DT and KM analyzed the data. DT conducted the literature review and wrote the first draft of the manuscript and MB had responsibility for final content. All authors read, contributed to and approved the final manuscript.

## Notes

***Financial Support Statement:*** This work was supported by a UWI Graduate Studies Research and Publications Grant.

***Conflict of Interest Statement:*** The authors have no conflicts to declare.

### Competing Interest Statement

The authors have declared no competing interest.

### Funding Statement

This study was funded by a a UWI Graduate Studies Research and Publications Grant.

### Author Declarations

The Mona Campus Research Ethics Committee of the University of the West Indies gave ethical approval for this work.

